# Genetic Testing History in Adults with Autism Spectrum Disorder

**DOI:** 10.1101/2024.08.18.24312179

**Authors:** Susanna B. Mierau, Robyn P. Thom, Caitlin T. Ravichandran, Amanda Nagy, Cashel Rice, Christina Macenski, Christopher J. Keary, Michelle L. Palumbo, Christopher J. McDougle, Ann M. Neumeyer

## Abstract

**Purpose:** Many genes have been identified in autism spectrum disorder (ASD). Yet how many adults with ASD receive recommended genetic testing and their outcomes is unknown. We investigated the percentage of adults with ASD with documented genetic testing in our ASD specialty clinic and the percentage with positive findings.

**Methods:** Adults were identified through search of our data repository and ASD diagnoses confirmed using record review by psychiatrists specializing in ASD. Patients were included (N=630) who had at least one visit with a qualifying clinician between 5/1/2010 and 12/15/2020. Data were collected through manual retrospective record review.

**Results:** Only 41% of the adults with ASD (261/630) had a documented history of genetic testing. Genetic testing was declined by patients or families for 11% of records and not recorded in 47%. Mean (SD; range) age for the 261 adults was 28.5 (5.3; 22-58) years; 26% were female and 73% had intellectual disability (ID). The genetic testing method was recorded in 91% (238). Only 54% of these patients had testing using a recommended method (chromosomal array, autism/ID sequencing panel, or exome sequencing). Few adults received testing with sequencing technologies. A genetic cause of ASD was found in 28%.

**Conclusion:** ASD-related genetic testing is underutilized in adults with ASD. Nearly half of the adults in our sample lacked documentation of genetic testing. Thus, the percentage who received testing may be even lower than reported. Adults with ASD may benefit from having their genetic testing history reviewed in the clinic and the recommended testing performed.

## Introduction

Autism spectrum disorder (ASD) is a lifelong condition that affects social communication and interaction and may lead to repetitive behaviors and reduced or heightened sensory sensitivities. The estimated prevalence of adults with ASD was 2.21% in the U.S. in 2017 (Dietz et al., 2020). The prevalence in adults will likely continue to increase as more people with ASD turn 18 years; the prevalence in children was 1 in 36 in 2020 (Maenner et al, 2023). With the increased availability of genetic testing and large family-based genetic studies, the number of known genes that cause ASD is increasing (Vorstman et al, 2017; Zhou et al, 2022; Wright et al, 2024). Knowledge of the genetic cause (or causes where multiple gene variants are identified) for the individual and family can have a positive effect on the understanding of the cause of lifelong difficulties (Martin et al, 2020; Wright et al, 2024; Klitzman et al, 2024; Wynn et al, 2025). The genetic cause can also provide important clinical information, particularly where other organ systems may be involved (Finucane et al, 2020). For patients with limited ability to communicate pain or discomfort in their body, this knowledge can be lifesaving (e.g., increased lifetime cancer risk with *PTEN* variants; Finucane et al, 2020). Support groups for individuals and family members with specific genetic causes of ASD are also forming at the national and international level (Wright et al, 2024). At the population level, identifying groups of people with a similar genetic cause of ASD may also facilitate the development of better therapeutic strategies, including more personalized medicine, and address the challenge of phenotypic heterogeneity in ASD clinical studies (Finucane et al, 2020).

Many adults for whom ASD was identified in childhood have had testing for Fragile X syndrome and a chromosomal microarray. In 2010, these two tests were recommended as first tier by the American College of Medical Genetics and Genomics (ACMG) for people with unexplained developmental delays, intellectual disability, and/or ASD (Miller et al, 2010). In the last 10 years, however, larger gene panels that test for thousands of ASD genes have become available that are commonly used in children. More recently, whole exome and now whole genome sequencing has also become available clinically. A 2019 consensus statement, based on the lower yield of chromosomal microarray compared to exome sequencing, recommended exome sequencing as a first-tier clinical diagnostic test for people with neurodevelopmental disorders, including autism spectrum disorder (Srivastava et al., 2019). The ACMG now recommends both exome and genome sequencing as first (or second) tier for children with congenital anomalies, developmental delay or intellectual disability with onset prior to 18 years of age (Manickam et al., 2021). Whole exome sequencing in an Indian cohort, for example, yielded a genetic diagnosis for ASD in 29.7% of 101 children versus 2.9% from chromosomal microarray (Sheth et al., 2023). Whole genome sequencing is increasing the diagnostic yield further, particularly through large population-level studies from individuals of primarily European (e.g., Trost et al., 2022) and East Asian ancestry (e.g., Kim et al., 2024).

These newer tests, combined with the increase in knowledge of gene variants that cause ASD, create an opportunity to improve clinical care. However, there is limited information available on what percentage of adults with ASD have received up-to-date genetic testing. Low rates of clinical ASD-related genetic testing were reported in a primarily pediatric cohort in Rhode Island with 132 individuals over the age of 20 (Moreno-De-Luca et al., 2020) and in Swedish cohort of 213 teens and adults (Hellquist and Tammimies, 2022). To address this gap, we investigated the percentage of 630 adults with confirmed ASD diagnosis who were seen in our clinic between 2010-2020 who had documentation of any of the recommended genetic testing methods (i.e., chromosomal array, autism/intellectual disability sequencing panel, or exome sequencing) and the percentage of positive findings for those who received genetic testing.

## Methods

### Ethics

This study was approved by the Mass General Brigham Institutional Review Board.

### Eligibility Criteria

This was a retrospective chart review study using a previously characterized patient cohort from our autism specialty clinic (Thom et al., 2022). In brief, eligibility criteria included: (1) past or current patient at the Massachusetts General Hospital (MGH) Lurie Center for Autism; (2) documented developmental history; (3) comprehensive clinical evaluation by a Lurie Center or other MGH developmental pediatrician, psychiatrist, psychologist, neuropsychologist, or neurologist; and (4) support for an ASD diagnosis. In the current study, patients were included who had at least one visit with a qualifying clinician from our hospital between 5/1/2010 and 12/15/2020.

### Information Sources

Potentially eligible adults were originally identified for an ongoing study investigating co-occurring medical conditions in adults with an ASD diagnosis (the “original” study). The identification process and confirmation of ASD diagnosis are described in a previous publication (Thom et al., 2022). Medical records from adults excluded from the original study due to genetic conditions were reviewed for ASD diagnosis to determine eligibility for the current study. Updated demographic and encounter history information for adults with a confirmed ASD diagnosis were obtained from the Mass General Brigham Research Patient Data Registry (RPDR).

For the current study, data were most recently requested from the RPDR on May 12, 2021. Data on the presence and severity of intellectual disability were available for a subset of adults included in a study of cardiometabolic risk factors (Thom et al., 2022). Further treatment history information and clinical and genetic testing data were collected using manual review of the electronic health record (EHR) from December 2020 to September 2022, with a medical record cutoff date of December 15, 2020.

### Selection Process

Figure 1 illustrates the study flow for the selection process of the original study and the current study. Most potentially eligible adults had a confirmed ASD diagnosis from the original study. For 33 patients who were ineligible for the original study due to genetic conditions, medical records pertaining to developmental and clinical history were reviewed by the same expert psychiatrists participating in the original study to confirm ASD diagnosis. For all patients with a confirmed ASD diagnosis, data from the Research Patient Data Registry (RPDR) and the patients’ electronic health records (EHRs) were reviewed for qualifying visits.

**Figure 1.**
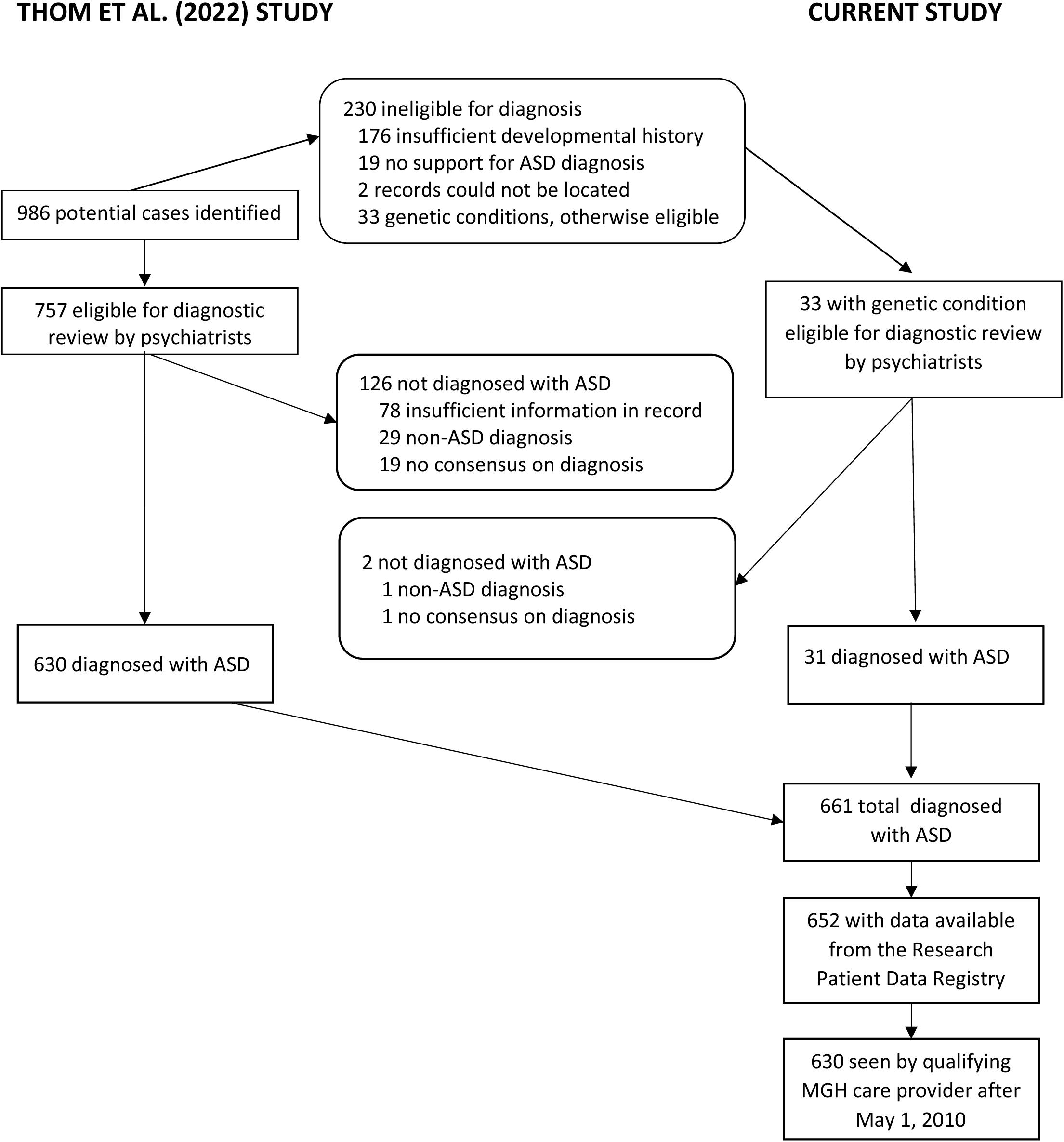
Study Flow. Diagram illustrates steps in the selection of cases for the current study with confirmed ASD diagnosis and the relationship to cases used in a previous study (Thom et al., 2022).

### Measures

Demographic characteristics of patients including age, sex, race, and ethnicity, were classified using data returned from the RPDR. Data on clinical and family history of the patient were coded based on retrospective manual review of developmental pediatrics, psychiatry, neurology, and primary care notes in the EHR. A keyword search of the full EHR for “seizure” was used to verify the presence or absence of seizure history. Presence or absence of intellectual disability was assigned based on full scale IQ, when documented in the EHR. If full scale IQ was not documented, presence or absence of intellectual disability was assigned based on qualitative assessment in the clinical notes or documentation of services through the Department of Developmental Services.

Genetic testing history was determined based on a keyword search of the full EHR for “genetic” and subsequent review of relevant clinical documentation and lab testing. For each occurrence of genetic testing, the date of sampling, type of test, and testing results were recorded. Testing type history was summarized over time as one or more tests using a recommended method (chromosomal microarray, autism ID panel, or exome sequencing), karyotype testing only, or other non-recommended method only. Testing results were classified as positive, negative, or variant of unknown significance (VUS) based on the lab report associated with the testing or, if lab results were unavailable in the EHR, documentation of testing results by a clinician. Final decisions on the categorization of type of testing and designation of testing as ASD-related were made by the study’s senior author.

### Statistical Approach

Demographic and clinical characteristics of adults in the sample were summarized by genetic testing history using means, standard deviations, and ranges for continuous variables and frequencies and percentages for categorical variables. Timing of earliest ASD clinic encounter and most recent ASD clinic encounter were categorized based on their observed distributions in the sample to achieve adequate distribution of adults across categories. Robust linear regression (continuous variables) and chi-square tests (categorical variables) assessed the statistical significance of differences in characteristics among those with testing information in the EHR, those without testing history in the EHR, and those who declined genetic testing. Ninety-five percent confidence intervals (CIs) for percentages were calculated using Wilson’s method for binomial confidence intervals (specific genetic tests) and Goodman’s method for multinomial confidence intervals (testing history summary, testing results; Goodman, 1965). Associations of candidate demographic and clinical predictors with frequency of testing using a recommended method among those adults with testing history documented were quantified using relative risk regression models fit with the modified Poisson approach (Zou, 2004). Due to the modest number of adults with a history of testing using a recommended method and the potential for strong associations between candidate predictors, associations with candidate clinical predictors were initially assessed individually in single predictor models controlling for age and sex. Clinical predictors associated with p-values less than 0.10 in the single predictor models were then entered together into a multiple predictor model, from which statistical significance was assessed.

Adults with missing values for demographic, clinical, and genetic testing variables were excluded from corresponding data analyses; details on frequency of missing data are provided in table and figure footnotes. Data analysis was conducted using version 9.4 of SAS (SAS Institute, Cary, NC) and version 4.3.2 of R (R Core Team, Vienna, Austria). Multinomial confidence intervals were calculated using the MultinomCI function from the R DescTools package (Signorell, 2024). All statistical tests were two-sided and conducted at the test-wise alpha=0.05 significance level.

### Data Availability

The authors will share the de-identified data used for this study with qualified researchers whose proposal of data use has been approved by an independent institutional review committee.

### Study Selection

The 630 adults with confirmed ASD diagnoses from the original study were combined with 31 adults, who although ASD diagnosis was confirmed were excluded from the original study because they had a known genetic diagnosis. Of the 661 patients who met the eligibility criteria, records for 652 patients were available in the research database. Six hundred thirty of these patients were seen by a qualifying provider from our hospital between May 1, 2010 and December 15, 2020, and thus were included in the current study.

### Study Characteristics

The demographic and clinical characteristics of the 630 adults included in this study are described in Table 1.

**Table 1:**
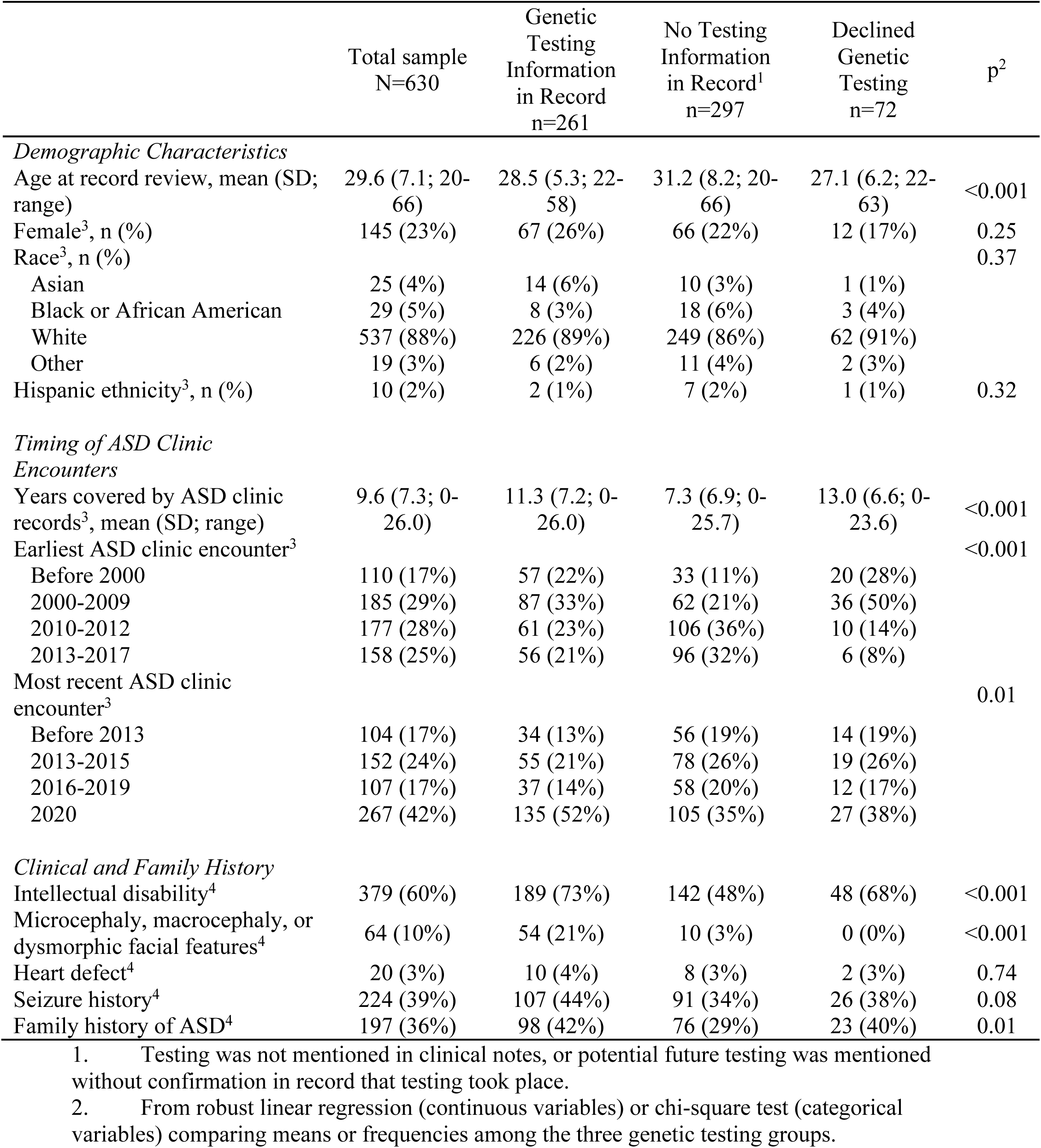
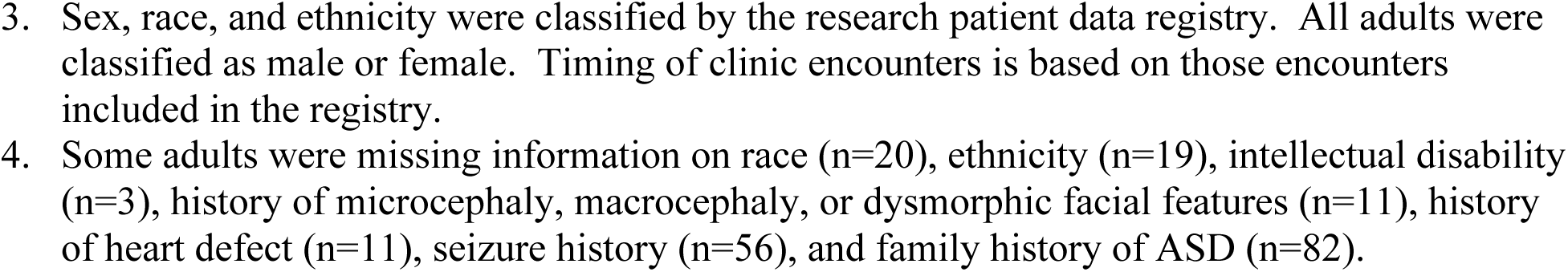
Demographic and Clinical Characteristics of the Adults by Availability of Genetic Testing History.

|  | Total sample<br>N=630 | Genetic<br>Testing<br>Information<br>in Record<br>n=261 | No Testing<br>Information<br>in Record<sup>1</sup><br>n=297 | Declined<br>Genetic<br>Testing<br>n=72 | p<sup>2</sup> |
| --- | --- | --- | --- | --- | --- |
| *Demographic Characteristics* |  |  |  |  |  |
| Age at record review, mean (SD; range) | 29.6 (7.1; 20-66) | 28.5 (5.3; 22-58) | 31.2 (8.2; 20-66) | 27.1 (6.2; 22-63) | <0.001 |
| Female<sup>3</sup>, n (%) | 145 (23%) | 67 (26%) | 66 (22%) | 12 (17%) | 0.25 |
| Race<sup>3</sup>, n (%) |  |  |  |  | 0.37 |
| Asian | 25 (4%) | 14 (6%) | 10 (3%) | 1 (1%) |  |
| Black or African American | 29 (5%) | 8 (3%) | 18 (6%) | 3 (4%) |  |
| White | 537 (88%) | 226 (89%) | 249 (86%) | 62 (91%) |  |
| Other | 19 (3%) | 6 (2%) | 11 (4%) | 2 (3%) |  |
| Hispanic ethnicity<sup>3</sup>, n (%) | 10 (2%) | 2 (1%) | 7 (2%) | 1 (1%) | 0.32 |
| *Timing of ASD Clinic Encounters* |  |  |  |  |  |
| Years covered by ASD clinic records<sup>3</sup>, mean (SD; range) | 9.6 (7.3; 0-26.0) | 11.3 (7.2; 0-26.0) | 7.3 (6.9; 0-25.7) | 13.0 (6.6; 0-23.6) | <0.001 |
| Earliest ASD clinic encounter<sup>3</sup> |  |  |  |  | <0.001 |
| Before 2000 | 110 (17%) | 57 (22%) | 33 (11%) | 20 (28%) |  |
| 2000-2009 | 185 (29%) | 87 (33%) | 62 (21%) | 36 (50%) |  |
| 2010-2012 | 177 (28%) | 61 (23%) | 106 (36%) | 10 (14%) |  |
| 2013-2017 | 158 (25%) | 56 (21%) | 96 (32%) | 6 (8%) |  |
| Most recent ASD clinic encounter<sup>3</sup> |  |  |  |  | 0.01 |
| Before 2013 | 104 (17%) | 34 (13%) | 56 (19%) | 14 (19%) |  |
| 2013-2015 | 152 (24%) | 55 (21%) | 78 (26%) | 19 (26%) |  |
| 2016-2019 | 107 (17%) | 37 (14%) | 58 (20%) | 12 (17%) |  |
| 2020 | 267 (42%) | 135 (52%) | 105 (35%) | 27 (38%) |  |
| *Clinical and Family History* |  |  |  |  |  |
| Intellectual disability<sup>4</sup> | 379 (60%) | 189 (73%) | 142 (48%) | 48 (68%) | <0.001 |
| Microcephaly, macrocephaly, or dysmorphic facial features<sup>4</sup> | 64 (10%) | 54 (21%) | 10 (3%) | 0 (0%) | <0.001 |
| Heart defect<sup>4</sup> | 20 (3%) | 10 (4%) | 8 (3%) | 2 (3%) | 0.74 |
| Seizure history<sup>4</sup> | 224 (39%) | 107 (44%) | 91 (34%) | 26 (38%) | 0.08 |
| Family history of ASD<sup>4</sup> | 197 (36%) | 98 (42%) | 76 (29%) | 23 (40%) | 0.01 |

1. Testing was not mentioned in clinical notes, or potential future testing was mentioned without confirmation in record that testing took place.
2. From robust linear regression (continuous variables) or chi-square test (categorical variables) comparing means or frequencies among the three genetic testing groups.
3. Sex, race, and ethnicity were classified by the research patient data registry. All adults were classified as male or female. Timing of clinic encounters is based on those encounters included in the registry. 4. Some adults were missing information on race (n=20), ethnicity (n=19), intellectual disability (n=3), history of microcephaly, macrocephaly, or dysmorphic facial features (n=11), history of heart defect (n=11), seizure history (n=56), and family history of ASD (n=82).

## Results

### Characteristics of the Adults and Documentation of Testing History

Only 41% of the adults with ASD (261/630) had a history of ASD-related genetic testing documented in the medical record. Genetic testing was declined by the patient or family for 11% (72) of records and not recorded in 47% (297). Among the 630 adults, 28% (158/566) of those with complete data on clinical history had no history of intellectual disability, seizure, microcephaly, macrocephaly, dysmorphic facial features, or heart defect; and, among these 158, 23% (36) had a history of testing documented, 19 (12%) had a history of declined testing, and 65% (103) had no testing history recorded. The study included adults aged 20- to 66-years-old (mean of 29.6, SD 7.1 years) with 23% women. The mean (SD; range) age for the 261 adults with testing documented was 28.5 (5.3; 22-58) years. Sixty-seven (26%) were identified as female, 14 (6%) as Asian, 8 (3%) as Black or African American, 226 (89%) as White, 6 (2%) as other race, and 2 (1%) as Hispanic. One hundred eighty-nine (73%) had intellectual disability. Thus, the results presented here represent a clinic population that is primarily White and male.

The years covered by ASD clinic records for the adults in this study ranged from less than 1 year to 26.0 years (mean 9.6, SD 7.3). Patients with no genetic testing information in the record had on average 4.0 fewer years covered by ASD clinic records than those with genetic testing information and 5.7 fewer years than those who declined genetic testing (p<0.001, 3-group comparison). Interestingly, adults for whom the offer of genetic testing was declined were most likely to have had their earliest ASD encounter prior to 2010 (p<0.001, 3-group comparison across 4 date categories). Genetic testing information also varied by year of the most recent clinic visit (p=0.01, 3-group comparison across 4 date categories).

### Type of Genetic Testing Among Adults with a Documented History

Ninety-one percent (238/261) of adults with genetic testing information in the EHR had the genetic testing method recorded for all tests. Table 2 summarizes information on type of genetic testing method and displays the frequency of the most common testing methods. Fifty-four percent (95% CI: 46%, 61%) of adults with testing information had been tested using a recommended method, 6% (95% CI: 3%, 11%) had been tested using karyotype only, and 40% (95% CI: 33%, 48%) had been tested using other non-recommended methods only. Older methods, including Fragile X testing, chromosomal microarray, and karyotype, were the most common methods documented. Newer methods such as an autism and/or intellectual disability sequencing gene panel and whole sequencing were identified in 7% (95% CI: 4%, 11%) and 7% (95% CI: 4%, 11%), respectively, of the adults with any history of genetic testing in the record. Among the 13% (32) of adults with genetic testing method recorded for all tests who had no clinical history of intellectual disability, seizure, microcephaly, macrocephaly, dysmorphic facial features, or heart defect, 50% (95% CI: 31%, 69%) had been tested using a recommended method, 9% (95% CI: 3%, 28%) had been tested using karyotype only, and 41% (95% CI: 23%, 61%) had been tested using other non-recommended methods only.

**Table 2:** Percentage with Genetic Testing History by Testing Type, n=238 Adults with Any History of ASD-related Genetic Testing in Record and Complete History of Testing Type^1^.

| Testing Type | Percentage<br>(95% Confidence<br>Interval) |
| --- | --- |
| *Summary* |  |
| Any recommended method<sup>2</sup> | 54% (46%, 61%) |
| Karyotype only | 6% (3%, 11%) |
| Other non-recommended method only | 40% (33%, 48%) |
| *Individual Tests* |  |
| Fragile X | 71% (65%, 76%) |
| Chromosomal microarray | 50% (44%, 57%) |
| Karyotype | 39% (33%, 45%) |
| Angelman syndrome or Prader Willi syndrome | 8% (5%, 12%) |
| Rett syndrome | 8% (5%, 12%) |
| Autism/Intellectual Disability panel | 7% (4%, 11%) |
| Exome sequencing | 7% (4%, 11%) |
| Mitochondrial DNA | 4% (2%, 8%) |
| Other single gene/targeted<sup>3</sup> | 11% (8%, 16%) |
| Other testing<sup>4</sup> | 1% (0%, 3%) |

1. Of 261 adults with genetic testing information in their electronic health records, 238 (91%) had information on test type for all tests mentioned in the record. One adult with a history of only non-ASD related testing is also excluded from calculations. 2. Chromosomal microarray, autism / intellectual disability (ID) gene panel, or exome sequencing 3. Unspecified FISH (n=11), PTEN (n=4), SCN1A (n=3), STK9/CDKL5 (n=3), Smith Magenis (n=2), MED12 (n=2), NLGN3 and NLGN4 (n=2), ARX (n=2), velocardiofacial syndrome (n=2), chromosome 15 (n=2), ATRX (n=1), 22q (n=1), SLC22A5 (n=1), RSK2 (n=1), SLC6A8 (n=1), DiGeorge (n=1), and CATCH 22 (n=1) 4. Chromosome 15 microsatellite (n=1) and mucopolysaccharidosis type III panel (n=1)

Of the 129 adults with a record of testing using a recommended method, 62 had no record of genetic testing prior to testing using the recommended method, 32 had a record of previous testing with a negative or VUS result, and 35 had an incomplete testing history. That is, the dates or outcomes of non-recommended testing were not available in the record.

Estimated frequency ratios associating demographic and clinical characteristics of patients with testing using a recommended method among the 238 adults with type of testing recorded are reported in Table 3. The presence of microcephaly, macrocephaly, or dysmorphic facial features was associated with 1.58 (95% CI: 1.27, 1.97) times the frequency of recommended testing after adjustment for age, sex, and seizure history (p<0.001), and seizure history was associated with 1.29 (95% CI: 1.01, 1.64) times the frequency of recommended testing after controlling for presence of microcephaly, macrocephaly, or dysmorphic facial features, age, and sex (p=0.04). Testing using a recommended method was not significantly associated with age, sex, intellectual disability, family history of ASD, or seizure history.

**Table 3:** Associations of Demographic and Clinical Characteristics with Genetic Testing Using a Recommended Method, Estimated Frequency Ratio (95% Confidence Interval), n=238 Adults with Any History of ASD-related Genetic Testing in Record and Complete Testing History.

|  | Single Predictor Model |  | Multiple Predictor Model |  |
| --- | --- | --- | --- | --- |
|  | Frequency Ratio (95% CI) | P | Frequency Ratio (95% CI) | P |
| Age (10 year increase) | 0.94 (0.74, 1.20) | 0.63 | 0.91 (0.73, 1.13) | 0.39 |
| Female | 0.97 (0.73, 1.28) | 0.82 | 1.00 (0.76, 1.33) | 0.98 |
| Intellectual disability | 0.96 (0.74, 1.24) | 0.75 |  |  |
| Family history of ASD | 1.04 (0.82, 1.33) | 0.72 |  |  |
| Seizure history | 1.40 (1.10, 1.78) | 0.006 | 1.29 (1.01, 1.64) | 0.04 |
| Microcephaly, macrocephaly, or dysmorphic facial features | 1.66 (1.34, 2.06) | <0.001 | 1.58 (1.27, 1.97) | <0.001 |

CI=confidence interval. Estimated frequency ratios and p-values are from relative risk regression models. All models included covariates for age at record review and sex. Characteristics associated with a p-value <0.10 in the single predictor model were entered together into a multiple predictor model. Data from adults with missing covariate values were excluded from corresponding regression models. Two adults were missing intellectual disability, 26 adults were missing family history of ASD, 18 adults were missing seizure history, and 3 adults were missing microcephaly, macrocephaly, or dysmorphic facial features. A total of 218 adults contributed data to the multiple predictor model.

### Results of Genetic Testing Using Recommended Methods

Current recommendations for genetic testing have changed from chromosomal microarray to whole exome sequencing and, in some cases, whole genome sequencing. For the time period covered in this study recommended testing included chromosomal microarray and, if unrevealing, an autism and/or intellectual disability gene panel, as whole exome sequencing was not yet available and/or covered by medical insurance during part of the time period covered in this study. During the time period where gene panels were in common use, exome sequencing may also have been performed if the gene panel was negative. The outcomes of ASD-related genetic testing using recommended methods, as a whole and for the three recommended testing methods, are provided in Table 4. For adults with a documented history of multiple tests using recommended methods, results are summarized across tests.

**Table 4:**
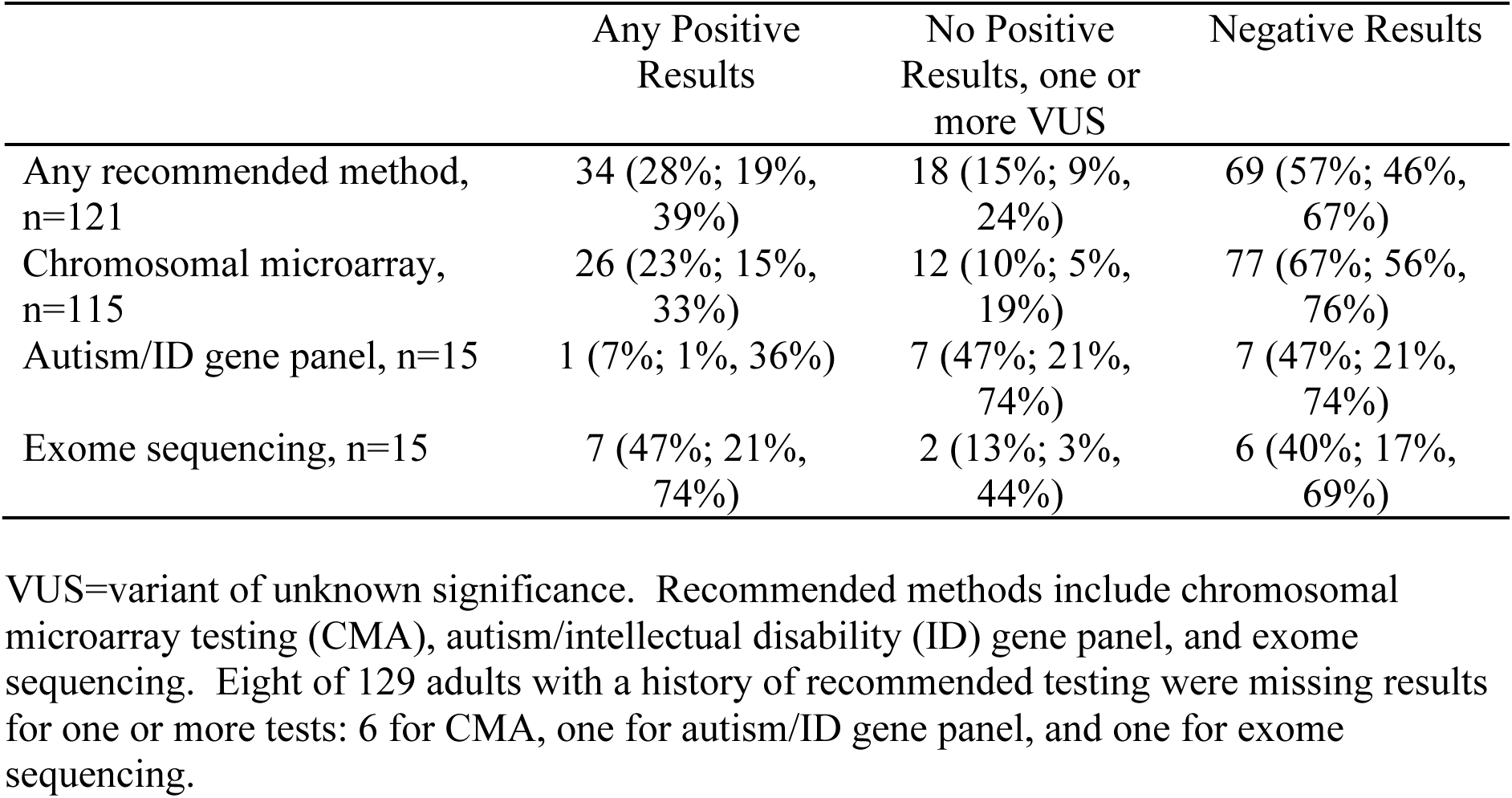
Outcomes of Genetic Testing Using Recommended Methods, n (%; 95% Confidence Interval)

|  | Any Positive Results | No Positive Results, one or more VUS | Negative Results |
| --- | --- | --- | --- |
| Any recommended method, n=121 | 34 (28%; 19%, 39%) | 18 (15%; 9%, 24%) | 69 (57%; 46%, 67%) |
| Chromosomal microarray, n=115 | 26 (23%; 15%, 33%) | 12 (10%; 5%, 19%) | 77 (67%; 56%, 76%) |
| Autism/ID gene panel, n=15 | 1 (7%; 1%, 36%) | 7 (47%; 21%, 74%) | 7 (47%; 21%, 74%) |
| Exome sequencing, n=15 | 7 (47%; 21%, 74%) | 2 (13%; 3%, 44%) | 6 (40%; 17%, 69%) |

VUS=variant of unknown significance. Recommended methods include chromosomal microarray testing (CMA), autism/intellectual disability (ID) gene panel, and exome sequencing. Eight of 129 adults with a history of recommended testing were missing results for one or more tests: 6 for CMA, one for autism/ID gene panel, and one for exome sequencing.

Notably, the identification of a genetic cause of ASD (labeled as “any positive results” in the table) was found in 28% (95% CI: 19%, 39%) of the patients who had one or more of the recommended methods for ASD-related genetic testing documented. The majority of the patients in this study with documentation of one or more recommended methods had a chromosomal microarray with 23% (95% CI: 15%, 33%) having a positive test. An autism/intellectual disability gene panel had 1 positive result in the 15 patients with reported testing; however, 47% (7/15; 95% CI: 21%, 74%) had one or more variant of unknown significant (VUS) identified. VUS describes changes in an ASD-related gene in which there is not yet enough information to determine if the specific genetic mutation is benign or pathogenic. Exome sequencing had 47% (7/15; 95% CI: 21%, 74%) positive results with an additional 13% (2/15; 95% CI:3%, 44%) with one or more VUS. Among 16 adults who had one or more of the recommended methods for ASD-related genetic testing documented, and who had no history of intellectual disability, seizure, microcephaly, macrocephaly, dysmorphic facial features, or heart defect, 25% (4/16; 95% CI: 8%, 55%) had positive results, 56% (9/16; 95% CI: 29%, 80%) had negative results, and 19% (3/16; 95% CI: 5%, 49%) had one or more VUS.

## Discussion

### Main Findings

This study shows that, even at a highly specialized ASD clinic, the percentage of adults with documented ASD-related genetic testing history in patients seen by a qualifying provider from 2010-2020 was only 41% (261 of 630 adults). Of the adults with documented ASD-related genetic testing history, only 54% (95% CI: 46%, 61%) had received one or more of the recommended methods (chromosomal microarray, autism/intellectual disability gene panel, or exome sequencing). Importantly, 28% (95% CI: 19%, 39%) of adults with documentation of one or more of the recommended methods received a genetic diagnosis with an additional 10% (95% CI: 5%, 19%) for whom new gene variants were identified that may cause ASD. Although fewer adults during this time period (2009-2020) were tested using an autism/intellectual disability gene panel or exome sequencing, pathogenic or unknown significance variants in ASD-related genes were identified in 53% (95% CI: 26%, 79%) with the gene panel and 60% (95% CI: 31%, 83%) with exome sequencing. Together, these findings highlight the potential benefit of reviewing genetic testing history in adults with ASD and performing recommended ASD-related genetic testing to identify, where possible, the genetic cause or causes of ASD.

### Relationship to Other Reports

There is very limited information in the medical literature on the percentage of adults with ASD who receive ASD-related genetic testing as part of their routine clinical care. A retrospective chart review study (2013-2019) of 1280 children and adults diagnosed with ASD based on the ADOS-2 reported that only 16.5% had received any ASD-related genetic testing (Moreno-De-Luca et al., 2020). Only 10% (132) of the patients were 20 years or older in this cohort from the Rhode Island Consortium for Autism Research and Treatment (RI-CART) study. The percentage of these adults with any genetic testing was very low: Fragile X (∼12%), karyotype (∼7%), and chromosomal microarray (∼2.5%). Interestingly, genetic testing was more likely to be ordered by pediatric subspecialists than psychiatrists or psychologists. This study is consistent with our findings in our larger population of adults with confirmed ASD that many adults lack the recommended ASD-related genetic testing. It also suggests that the percentage of adults with ASD-related genetic testing may be much lower in the community than in ASD specialty clinics.

A 2021 study in Sweden found that only 2.8% of the 213 adolescents and adults had received a referral for genetic testing after an ASD diagnosis (Hellquist and Tammimies, 2022). Interestingly, the prevalence of ASD in 18-24 years (older ages not reported here) was 2.4%, similar to the percentage of adults with ASD in Massachusetts, and the recommendation in Sweden by several medical societies, similar to here in the U.S., is that clinical genetic testing be offered to everyone with ASD. Recommendations in Sweden included chromosomal microarray and exome sequencing. Whether autism/intellectual disability gene panels, that have been commonly used in American clinical genetic testing, were used was not mentioned. There are multiple reviews advocating for adults with ASD to receive recommended ASD-related genetic testing (Schaefer, 2016; Kreiman and Boles, 2020; Fiucane et al, 2020) consistent with our experience that many adults still need to be offered ASD-related genetic testing. In our study, 28% of patients or families who were first seen at our clinic prior to 2010 had declined genetic testing when offered, although this percentage decreased for patients who joined the clinic after 2010. Surveys of autistic individuals (Byres et al., 2023; Gallion et al., 2024; Klitzman et al, 2024b) and the parents of children with ASD (Zhou et al., 2023; Klitzman et al, 2024a) report a range of opinions with some favoring autism-specific genetic testing while others opposing. Interestingly, adults who received prior genetic testing had more positive beliefs about autism-related genetic testing compared to those without prior testing (Gallion et al., 2024) and, in an international meta-analysis, 50-59.6% of parents who previously obtained genetic testing would recommend it to other parents (Zhou et al., 2023).

A recent meta-analysis by leading investigators in the field of neurodevelopmental (NDD) genetics on the outcomes of genetic testing found that the yield of exome sequencing was 30-43% compared to 15-20% with chromosomal microarray (Srivastava et al., 2019). In isolated NDD, the yield was 31% (CI: 25-38%) and in NDD plus other conditions was 53% (CI: 41-64%), for example ASD with ID. The percentage of our patients with a positive result on chromosomal microarray was much higher than reported in this meta-analysis. This could be due to differences in the diagnoses included in the studies reviewed (and method for confirming the diagnosis); our study included only adults with an expert confirmed ASD diagnosis. It could also be that positive results on chromosomal microarray were more likely to be recorded in the medical record of adults with autism seen in the qualifying time period for our study (2010-2020). The yield of positive results on the exome sequencing in this study was consistent with our study (47%). This increases the confidence in the reliability of our results for exome sequencing, given that the number of patients with exome sequencing in our study was very small (15 adults).

### Limitations

This study highlights the need to review and document ASD-related genetic testing in adult patients. This study has the advantage of including a large cohort (630 adults) that range in age from 20- to 66-year-olds, as the majority of studies in ASD focus on ages 22 and younger despite the fact that most people with ASD are adults. However, the mean age in the study was 29.6 (SD 7.1); thus, the percentage with genetic testing and/or documentation could be even lower in older adults with ASD. Although autism prevalence is lower among non-Hispanic White children than other racial and ethnic groups (Maenner et al., 2020), our patient cohort has the limitation of being primarily White (88%) and non-Hispanic (98%). It is possible that the rates of ASD-related genetic testing performed and documented may be even lower in the general population than the population represented in our clinic’s cohort.

Importantly, all the patient records in this study were reviewed by multiple psychiatrists specialized in working with adults with ASD in our clinic, and the ASD diagnosis was confirmed by consensus. However, there were patients potentially eligible for our study for whom the records were insufficient to confirm the diagnosis of ASD (Figure 1). Thus, we do not know what percentage of the patients with insufficient records had an accurate ASD diagnosis and were excluded from the study. Notably, there are only 26% women included in this study. This 3:1 ratio of males to females with ASD, compared to the often cited 4:1 ratio, highlights an improvement in the inclusion of women with ASD in this study. However, community studies here in the Boston area highlight that the true male-to-female ratio may be closer to 1.8:1 men to women with ASD due to the delayed age of diagnosis for many girls and women with ASD and well-documented gender bias inherent to common diagnostic and research tools such as the Autism Diagnostic Observation Scale (ADOS; D’Mello et al., 2022). In a survey-base study, 11% of adults who responded to the survey self-reported prior genetic testing specifically for autism (Gallion et al., 2024). The majority had this testing as part of a research study (e.g., SPARK for Autism). Interestingly, the participants were 53.8% women, 75% White, and 53.2% had at least one autistic family member. 74% of participants had received a clinical diagnosis, while 26% were self-diagnosed. In contrast, our ASD specialty clinic serves a range of ASD severity levels. In particular, we see a larger number of moderately-to-severely affected adults with ASD (Level 2 and 3) because of the difficulties they often face receiving medical care in other, particularly adult, clinical settings. Our specialty clinic is designed to accommodate the special sensory and safety needs of these patients and is staffed, including receptionists and medical assistants, by people sensitive to needs of children and adults with severe forms of ASD.

There is also a risk of reporting biases due to data not covered in the EHR. We are unable to determine if adults without documentation of testing in their records never received testing or if they received testing that was undocumented. We also relied on the accuracy of clinical characterization documented in provider notes, and on the completeness and accuracy of genetic testing data available in the record. Despite these limitations, our findings that many adults lack the recommended ASD-related genetic testing, which could reveal a genetic cause (or causes), is still valid and highly relevant for clinicians caring for adults with ASD.

The time period of the record reviews is also relevant. We included patients who were seen between 2010 to 2020 by a qualified specialist in our hospital. Since 2020 (the latest date in this record review), the use of autism/intellectual disability gene panels became more common due to better insurance coverage (including for the majority of our patients who receive state-funded health insurance), which as of mid-2024 has now been replaced with whole exome sequencing. Moreover, the likelihood of receiving a genetic diagnosis, particularly for our moderate-to-severely affected adults with ASD (Level 2 and 3), is likely higher than reported here due to the increasing number of confirmed ASD-related genes included in the panel (from hundreds to thousands of genes). Previously categorized VUS have also been recategorized as pathogenic based on the findings of large population-level ASD-related genetic studies such as SPARK for Autism (Wang et al, 2022; Zhou et al, 2022). As of 2023, access to whole exome sequencing (WES) is also increasing with many insurance companies now covering the costs and is now the standard of care. The diagnostic yield—for autistic adults with and without co-occurring conditions such as ID or epilepsy—has and will likely continue to increase due to SPARK for Autism (Zhou et al, 2022) and other large genetic studies in autism worldwide. Notably, the autism/intellectual disability gene panels and WES were most likely to identify monogenic (single gene variant) causes of ASD. While the available clinical testing would report if multiple pathogenic variants (or VUS) were detected, it did not provide results on other possible polygenic causes of ASD, in which a combination of variants in multiple genes is necessary to cause ASD. Thus, the percentage of patients with monogenic or polygenic causes of ASD is likely higher than the percentage of patients who received positive results in our study, due to these limitations in the clinical genetic testing available at the time.

### Implications

Identification of the genetic cause(s) of ASD in adults can provide a powerful tool for improving quality and access to health care and other ASD-related services (Martin et al, 2020; Fiucane et al, 2020). For clinicians, the gene variant(s) provides critical information about prognosis and associated medical problems (Ficune et al, 2020). For adults with ASD, the genetic cause informs care and facilitates connection to support groups for specific genetic conditions (Ficune et al, 2020; Klitzman et al, 2024b; Wright et al, 2024). For researchers, the genetic variants aid mechanistic and novel therapeutic studies. Our study reveals that many adults with ASD are likely yet to have recommended genetic testing. As ASD-related genetic testing is now more widely available and the number of ASD-related genes is expanding (Zhou et al, 2022), there is an opportunity now for clinicians to offer ASD-related genetic testing to their adult patients with ASD and for adults with ASD to receive the benefits of identifying the genetic cause (Fiucane et al, 2020; Wright et al, 2024). While insurance coverage may limit access to whole exome (or genome) sequencing for ASD-related genetic testing for adults without certain co-occurring conditions (e.g., ID, epilepsy, craniofacial anomalies), large ASD genetic studies, such as SPARK for Autism, are returning genetic diagnoses to autistic adults without co-occurring conditions (Wright et al, 2024) and expanding the list of ASD-causing gene variants in autistic individuals with and without related co-occurring conditions (Zhou et al, 2022).

Notably, there may be an education gap for patients as well as providers when considering ASD-related genetic testing. A Canadian survey of 461 autistic individuals found that only 27% would have wanted genetic testing during childhood and only 35% felt that testing should be routinely offered to autistic adults (Byres et al., 2023). Here we, along with others, advocate for increasing access to and documentation of genetic testing for adults with ASD (Fiucane et al, 2020; Wright et al, 2024). The SPARK for Autism study (over 100,000 participants) and other large genetic studies reveal that genetic diagnoses can be provided to and have a positive impact for adults with autism, including adults with no co-occurring conditions (e.g., ID, epilepsy, craniofacial anomalies) that would have provided insurance coverage for clinical genetic testing (Wright et al, 2024; Klitzman et al, 2024b). There is a concern among many adults who identify as autistic that ASD-related genetic testing could lead to a negative selection for embryos, for example, that have an ASD-related gene (Klitzman et al, 2024b). It is important to distinguish this concern from the current study. All of the patients in this study are adults who have a confirmed diagnosis of autism spectrum disorder.

Knowledge of gene variants that cause ASD can inform medical care, particularly for patients in which the genetic variant (or variants) affects multiple organ systems (Kreiman and Boles, 2020; Fiucane et al, 2020). This can be lifesaving for adults with language impairment and/or reduced ability to report pain or discomfort (Fiucane et al, 2020). Moreover, in multiple cases in our clinic, identification of a genetic cause lifts decades of guilt parents have carried that something they did (or did not do) during pregnancy, or at the time of birth, led to the severe ASD-related challenges their adult son or daughter faces. This has also been reported by others (Martin et al, 2020; Wright et al, 2024; Wynn et al., 2025). Identification of ASD-related gene variants has also enabled multiple women with ASD and co-occurring ID in our clinic to receive specialized services for ASD from the government, where the ASD diagnosis was not made until adulthood because of widespread and longstanding gender bias in ASD diagnosis.

## Conclusions

Ensuring that all adults with ASD are offered recommended genetic testing also has the potential to improve ASD clinical care at the individual and population level (Fiucane et al, 2020; Wright et al, 2024). Identifying the genetic cause(s) of ASD in an individual may enable personalized medicine (Schaefer 2016; Fiucane et al, 2020), such as ASD secondary to specific pathogenic variants in *MECP2* (Rett syndrome) that now have a specific therapy, trofinetide, that improves communication (Neul et al., 2024). By identifying groups of people with ASD who share the same genetic cause, future clinical and community-based studies can target interventions more precisely and reduce the phenotypic heterogeneity that has limited the ability of many ASD-related studies to show efficacy (Schaefer 2016; Wright et al, 2020).

## Author contributions

S.B.M. Major role in the interpretation of the data; drafting/revising the manuscript for content.

R.P.T. Acquisition of data; revising the manuscript for content.

C.T.R. Major role in the analysis and interpretation of the data; drafting/revising the manuscript for content.

A.N., C.R., C.M., C.J.K., M.L.P., C.J.M. Acquisition of data; revising the manuscript for content.

A.M.N. Major role in the study concept or design; interpretation of the data; revising of the manuscript for content; supervision.

## References

Byres, L., Morris, E., & Austin, J. (2023). Exploring Autistic adults’ perspectives on genetic testing for autism. Genet Med, 25, 100021.

D’Mello, A. M., Frosch, I. R., Li, C. E., Cardinaux, A. L., & Gabrieli, J. D. E. (2022). Exclusion of females in autism research: Empirical evidence for a “leaky” recruitment-to-research pipeline. Autism Res, 15, 1929–1940.

Dietz, P. M., Rose, C. E., McArthur, D., & Maenner, M. (2020). National and State Estimates of Adults with Autism Spectrum Disorder. J Autism Dev Disord, 50, 4258–4266.

Finucane, B. M., Myers, S. M., Martin, C. L., & Ledbetter, D. H. (2020). Long overdue: including adults with brain disorders in precision health initiatives. Current opinion in genetics & development, 65, 47–52.

Gallion, T., Williams, Z. J., Niarchou, M., Duncan, L., Hooker, G., & Taylor, K. A. (2024). Attitudes of autistic adults toward genetic testing for autism. J Genet Couns. Epub 2024 May 25.

Goodman, L. A. (1965). On simultaneous confidence intervals for multinomial proportions. Technometrics, 7, 247–254.

Hellquist, A., & Tammimies, K. (2022). Access, utilization, and awareness for clinical genetic testing in autism spectrum disorder in Sweden: A survey study. Autism, 26, 1795–1804.

Kim, S.-W., Lee, H., Song, D. Y., et al. (2024). Whole genome sequencing analysis identifies sex differences of familial pattern contributing to phenotypic diversity in autism. Genome Med, 16, 114.

Klitzman, R., Bezborodko, E., Chung, W. K., & Appelbaum, P. S. (2024a). Parents’ views of benefits and limitations of receiving genetic diagnoses for their offspring. Child Care Health Dev, 50, e13212.

Klitzman, R., Bezborodko, E., Chung, W. K., & Appelbaum, P. S. (2024b). Views of Genetic Testing for Autism Among Autism Self-Advocates: A Qualitative Study. AJOB Empir Bioeth, 15, 262–279.

Kreiman, B. L., & Boles, R. G. (2020). State of the art of genetic testing for patients with autism: A practical guide for clinicians. Semin Pediatr Neurol, 34, 100804.

Maenner, M. J., Warren, Z., Williams, A. R., et al. (2023). Prevalence and characteristics of Autism Spectrum Disorder among children aged 8 years — Autism and developmental disabilities monitoring network, 11 sites, united states, 2020. MMWR Surveill Summ, 72, 1–14.

Manickam, K., McClain, M. R., Demmer, L. A., et al. (2021). Exome and genome sequencing for pediatric patients with congenital anomalies or intellectual disability: an evidence-based clinical guideline of the American College of Medical Genetics and Genomics (ACMG). Genet Med, 23, 2029–2037.

Martin, C. L., Wain, K. E., Oetjens, M. T., Tolwinski, K., Palen, E., Hare-Harris, A., Habegger, L., Maxwell, E. K., Reid, J. G., Walsh, L. K., Myers, S. M., & Ledbetter, D. H. (2020). Identification of Neuropsychiatric Copy Number Variants in a Health Care System Population. JAMA psychiatry, 77, 1276–1285.

Miller, D. T., Adam, M. P., Aradhya, S., et al. (2010). Consensus statement: chromosomal microarray is a first-tier clinical diagnostic test for individuals with developmental disabilities or congenital anomalies. Am J Hum Genet, 86, 749–764.

Moreno-De-Luca, D., Kavanaugh, B. C., Best, C. R., Sheinkopf, S. J., Phornphutkul, C., & Morrow, E. M. (2020). Clinical Genetic Testing in Autism Spectrum Disorder in a Large Community-Based Population Sample. JAMA Psychiatry, 77, 979–981.

Neul, J. L., Percy, A. K., Benke, T. A., et al. (2024). Trofinetide treatment demonstrates a benefit over placebo for the ability to communicate in rett syndrome. Pediatr Neurol, 152, 63–72.

Schaefer, G. B. (2016). Clinical genetic aspects of ASD spectrum disorders. Int J Mol Sci, 17.

Sheth, F., Shah, J., Jain, D., et al. (2023). Comparative yield of molecular diagnostic algorithms for autism spectrum disorder diagnosis in India: evidence supporting whole exome sequencing as first tier test. BMC Neurol, 23, 292.

Signorell, A. (2024). DescTools:Tools for Descriptive Statistics. R package version 09954. Epub 2024.

Srivastava, S., Love-Nichols, J. A., Dies, K. A., et al. (2019). Meta-analysis and multidisciplinary consensus statement: exome sequencing is a first-tier clinical diagnostic test for individuals with neurodevelopmental disorders. Genet Med, 21, 2413–2421.

Thom, R. P., Palumbo, M. L., Keary, C. J., Hooker, J. M., McDougle, C. J., & Ravichandran, C. T. (2022). Prevalence and factors associated with overweight, obesity, and hypertension in a large clinical sample of adults with autism spectrum disorder. Sci Rep, 12, 9737.

Trost, B., Thiruvahindrapuram, B., Chan, A. J. S., et al. (2022). Genomic architecture of autism from comprehensive whole-genome sequence annotation. Cell, 185, 4409–4427.e18.

Vorstman, J. A. S., Parr, J. R., Moreno-De-Luca, D., Anney, R. J. L., Nurnberger, J. I., & Hallmayer, J. F. (2017). Autism genetics: opportunities and challenges for clinical translation. Nat Rev Genet, 18, 362–376.

Wang, T., Kim, C. N., Bakken, T. E., et al. (2022). Integrated gene analyses of de novo variants from 46,612 trios with autism and developmental disorders. Proc Natl Acad Sci USA, 119, e2203491119.

Wright, J. R., Astrovskaya, I., Barns, S. D., Goler, A., Zhou, X., Shu, C., Snyder, L. G., Han, B., SPARK Consortium, Shen, Y., Volfovsky, N., Hall, J. B., Feliciano, P., & Chung, W. K. (2024). Return of genetic research results in 21,532 individuals with autism. Genet Med, 26, 101202.

Wynn, J., Karlsen, A., Huber, B., Levine, A., Salem, A., White, L. C., Luby, M., Bezborodko, E., Xiao, S., Chung, W. K., Klitzman, R. L., & Appelbaum, P. S. (2025). Impact of a Genetic Diagnosis for a Child’s Autism on Parental Perceptions. J Autism Dev Disord, 55, 1809–1823.

Zhou, M., Zhang, Y.-M., & Li, T. (2023). Knowledge, attitudes and experiences of genetic testing for autism spectrum disorders among caregivers, patients, and health providers: A systematic review. World J Psychiatry, 13, 247–261.

Zhou, X., Feliciano, P., Shu, C., Wang, T., Astrovskaya, I., Hall, J. B., Obiajulu, J. U., Wright, J. R., Murali, S. C., Xu, S. X., Brueggeman, L., Thomas, T. R., Marchenko, O., Fleisch, C., Barns, S. D., Snyder, L. G., Han, B., Chang, T. S., Turner, T. N., Harvey, W. T., … Chung, W. K. (2022). Integrating de novo and inherited variants in 42,607 autism cases identifies mutations in new moderate-risk genes. Nature Genetics, 54, 1305–1319.

Zou, G. (2004). A modified poisson regression approach to prospective studies with binary data. Am J Epidemiol, 159, 702–706.

